# WORLDWIDE CASE FATALITY RATIO OF COVID-19 OVER TIME

**DOI:** 10.1101/2020.10.04.20206599

**Authors:** Rohan Chaubal, Sadhana Kannan, Navin Khattry, Sudeep Gupta

**Author notes:** Corresponding Author: Dr Sudeep Gupta, MD, DM, Professor of Medical Oncology, Room 1109, 11^th^ Floor, HBB, Tata Memorial Centre, Mumbai-400012. These authors have contributed equally to this work (Rohan Chaubal, Sudeep Gupta). Ethics approval and consent to participate: Institutional ethics committee approval was not sought for this analysis, as per institutional guidelines, because it did not involve collection or handling of individual patient data. Availability of data and materials: The raw data used in this analysis is available in supplementary files. Competing interests: All authors declare no support from any organisation for the submitted work, no financial relationships with any organisations that might have an interest in the submitted work in the previous three years, and no other relationships or activities that could appear to have influenced the submitted work. Funding: There was no funding for this study. Authors’ contributions: Rohan Chaubal - conceptualization, data collection, data analysis, writing original draft, writing review & editing, Sadhana Kannan - data analysis, writing review & editing Navin Khattry - data analysis, writing review & editing, Sudeep Gupta – conceptualization, data analysis, supervision, writing original draft, writing review & editing.

## Abstract

**Background:** The case fatality ratio (CFR) of coronavirus disease 2019 (COVID-19) has been reported to be variable among different countries and regions but few analyses have tracked this ratio worldwide over time.

**Methods:** The primary objective was to assess the time-course evolution of CFR of COVID-19 in all countries with available data and secondary objective was to evaluate associations between country-wise CFR and country-level health, human development, demographic and economic parameters. Day-wise data of COVID-19 cases and deaths for each country was extracted from a public repository and countries with at least 1000 cases on cutoff date were clustered by unsupervised k-means on the basis of deaths per 100000 population (DP100K). Day-wise CFR (cumulative deaths divided by cumulative cases, multiplied by 100) for each country and cluster (country group) was plotted as time-series and country-level parameters were tested for association with CFR using weighted multiple linear regression.

**Results:** On September 24, 2020 there were 32140504 cumulative COVID-19 cases and 981792 deaths reported from 184 countries for a worldwide CFR of 3.06 % (95%CI 3.05 -3.07). Unsupervised k-means clustering in 157 countries with at least 1000 reported cases resulted in Clusters (country groups) A, B, C, D and E with centroid DP100K and CFR of 0.100 and 2.51 (95% CI 2.42-2.61), 0.503 and 2.28 (95% CI 2.23-2.33), 1.816 and 1.73 (95% CI 1.71-1.75), 7.395 and 1.76 (95% CI 1.75-1.76), and 36.303 and 3.82 (95% CI 3.82-3.83), respectively. In a log-log analysis DP100K and CFR were significantly positively correlated (R=0.3570, p<0.001) with each other. All country groups and majority of included countries showed a pattern of gradually increasing CFR from the beginning of pandemic, followed by a plateau and then a steady decline in CFR. Among 10 country-level parameters, GDP per capita (β=-0.483, p=0.000), hospital beds per population (β=-0.372, p<0.001), mortality from air pollution (β=-0.487, p=0.003) and population density (β=-0.570, p< 0.000) were significantly negatively associated while maternal mortality ratio (β=0.431, p=0.000) and age (β=0.635, p<0.000) were positively associated with CFR.

**Conclusions:** The CFR of COVID-19 has gradually increased over time in majority of countries at various stages of the pandemic, followed by a plateau and a steady decline. Population level COVID-19 mortality burden and CFR are significantly positively associated with each other.

## BACKGROUND

The novel coronavirus SARS-CoV-2, causative agent of coronavirus disease 2019 (COVID-19), has caused a worldwide pandemic of massive proportions with a large number of fatalities. There is no proven vaccine to prevent this infection and, as yet, no proven treatment with high efficacy. It is also uncertain whether infected individuals will develop long lasting immunity. SARS-CoV-2 causes severe disease, manifested primarily by pneumonia and respiratory failure, in a small proportion of cases and this is the primary cause of mortality. Because of the high person-to-person transmission, SARS-CoV-2 has overwhelmed the healthcare systems of many countries, including several high-income countries.

From a public health perspective, it is of great importance to be able to have accurate estimates of the case fatality ratio of an epidemic disease. The reported case fatality ratios of COVID-19 have ranged from 0.5% to almost 10% in various countries with reported variation within countries as well. The propensity to suffer severe disease and death from COVID-19 has been reported to be impacted by many factors including patient’s age and comorbidity status, with older individuals or those with comorbidities having a higher mortality rate [1-7].

Because these and other as yet unknown factors impact mortality and are variably distributed between countries and regions, it is difficult to compare unadjusted CFR across countries. However, within the same country taken as a unit, the variability in these factors is likely to be less, and therefore it may be relevant and insightful to track the unadjusted CFR within countries over time. Such an analysis could provide time trends that may help inform public health planning, forecasting and resource allocation.

We performed, and report here, an analysis of the day-wise case fatality ratio of COVID-19 in all countries, using an open source data repository that is regularly updated, starting from the first reported case in that repository, until the data cut-off date.

## METHODS

### Ethics Declaration

This analysis was performed by academic researchers from a large tertiary cancer centre in India. Institutional ethics committee approval was not sought for this analysis, as per institutional guidelines, because it did not involve collection or handling of individual patient data. All the raw data used in this analysis is available as supplementary files

### Objectives

The primary objective was to analyse the time-course evolution of COVID-19 CFR in individual countries, keeping a 24-hour day as the unit of time. The secondary objectives were to correlate the country-wise CFR with 10 country-level health, human development, demographic and economic parameters (listed below under ‘Data Extraction’) to identify any associations.

### Eligibility Criteria

Countries whose time sequential data for COVID-19, in terms of cumulative cases, cumulative deaths and cumulative recoveries, were reported in the COVID-19 GitHub Repository for the COVID-19 dashboard by the Center for Systems Science and Engineering of the Johns Hopkins University School of Medicine (CSSE-JHU) [8], were included in the analysis. We excluded data from two cruise ships (The Diamond Princess and MD Zandaam) which reported localized outbreaks of COVID-19. Data from countries with at least 1000 cumulative confirmed COVID-19 cases reported on the cut-off date were included for unsupervised clustering based on COVID-19 deaths per 100000 population (DP100K), plotting the CFR time-trends, and performing the weighted multiple linear regression between country-level parameters and CFR. The sources of data in this Repository include World Health Organization, various national governments and other publically accessible resources [9]. The analysis included data from the date of first reporting on this Repository until the data cut-off date.

### Data Extraction

Data on COVID-19 cumulative cases and deaths as on cut-off date were downloaded from the online Repository (Supplementary Table 1) (9). The downloaded data included the cumulative number of COVID-19 cases and deaths at 12 AM GMT on each date for each country for the included time period. Where reported, the data for different geographical regions or sovereign territories for a country was added up with data from its mainland territory to derive country-level estimates.

The latest available data on ten different indices of health [(% mortality from cardio vascular diseases (CVD), cancer, diabetes or chronic respiratory disease (CRD) in the age group 30-70, total smoking prevalence in age >/= 15 years, maternal mortality ratio per 100000 live births, infant mortality ratio per 1000 live births), human development (age standardized mortality rate per 100000 population attributed to household and ambient air pollution, physicians per 1000 population, and hospital beds per 1000 population), population demographics (median age of population, population density in persons per square kilometre), and economic development [(gross domestic product (GDP) per capita)] were extracted from the World Bank Open Data (health, human development and economic development indices) [10] and United Nations data hub (population demographics) (Supplementary Table 2) on September 1, 2020 [11]. Additionally, each country’s population for the year 2019 reported as of July 1, 2020 was also retrieved from the World Bank Open Data hub [10] for calculating death per 100,000 (DP100K) of the population for each country.

### Data analysis

Data analysis was performed using custom python scripts, GraphPad Prism version 8.02 for Windows, GraphPad Software, La Jolla California USA and Microsoft Excel (2019), Microsoft Corporation, Redmond, USA. Data visualization was performed using Tableau Software v 2020.3, Seattle, USA. The Case Fatality Ratio (CFR) was calculated as follows: 

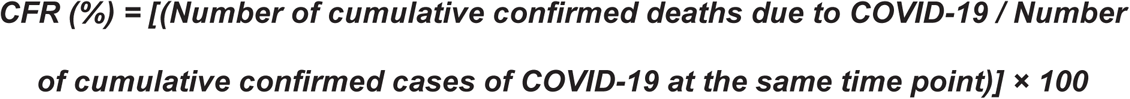

We calculated the COVID-19 deaths per 100000 population (DP100K) for each country using the total COVID-19 deaths on the cutoff date and country population for the year 2019. The DP100K was used to group countries with various levels of population-level COVID-19 mortality burden for depiction of CFR time-trends. Log (base 2) transformed DP100K on the x-axis was plotted against log (base 2) transformed CFR on the y-axis for countries with at least 1000 COVID-19 cases on the cutoff date and a linear regression, with coefficient of correlation, computed between these two variables. An unsupervised k-means clustering was performed based on log DP100K using Tableau, which uses Lloyd’s algorithm with squared Euclidean distances to estimate distances of all points in a cluster from the centroid of that cluster and centroids in other clusters. The algorithm estimates the sums of ‘between cluster’ and ‘within cluster’ distances which are used to calculate the ratio of variances for the above two metrics. At least 25 iterations were performed for ***k***=1…25 wherein for each value of ***k***, the ratio of the two variances was used to identify the first local maximum value for Calinski-Harabasz index (CH index) for smallest value of ***k***. We additionally generated Elbow plots to manually identify the appropriate ‘***k***’ for clustering and see its agreement with C-H generated ***k***. Wilcoxon signed rank was used to compare the median CFR of each country group identified by ***k***-means clustering and this data visualised as violin plots. We calculated the CFR for each day for each country from the downloaded data and depicted it date-wise for each DP100K cluster. Data points that were outliers and skewed visual representation were either plotted on a secondary y-axis and indicated in the graphs or removed from visualization (Supplementary Table 3).

We estimated the mean change in CFR per day for the most recent 10 calendar days from the cut-off date (including the cut-off date) for whole world, countries with >/= 1000 cases, country clusters, and each country, by subtracting the CFR on day minus10 from the CFR on the cut-off date, and dividing the difference by 10. A 2-sided 95% confidence interval for CFR on the cut-off date was calculated for the whole world, countries with >/= 1000 cases, each country cluster and each country. CFR for countries with >/= 1000 cases on the cut-off date were evaluated for independent association with 10 different country-level indices of health, human development, population demographics, economic development and mobility using weighted multiple linear regression analysis. Weighted regression was used because different countries with different numbers of cases and deaths contributed unequal amount of information. Weights were assigned using the inverse of the square of the standard error of CFR for each country. The multicollinearity assumption was tested using Variance Inflation Factor (VIF). Independence of residuals and homoscedasticity assumptions were checked using Durbin-Watson test and residuals plot (scatter plot of standardised residuals versus standardised predicted values), respectively. Variables which showed high collinearity (VIF > 10) were removed systematically at each step until the model only included factors which showed VIF < 10.

## RESULTS

### Case Fatality Ratio of COVID-19 over time

The data cut-off date was September 24, 2020 and included all cases and deaths due to COVID-19 in the CSSE-JHU Repository from the first date of its reporting, January 22, 2020. Of 195 countries, 184 have reported at least one confirmed case of COVID-19 (Figure 1, supplementary table 4) and are including in the analysis. In the 10 days preceding the cutoff date, 50 (27%) countries had a positive mean change in CFR, 118 (64%) had a negative mean change in CFR, while 16 countries (9%) had no net change in CFR (supplementary table 4). Figure 2.A depicts a map of the world colour coded with different ranges of CFR as on September 24, 2020, which shows that developed as well as less developed countries show high and low CFR. One hundred fifty-seven countries with case load >/= 1000 with 32133634 cumulative cases and 981664 cumulative deaths are included in the CFR time-trend analysis, unsupervised clustering analysis, and the weighted multiple linear regression analysis.

**Figure 1:**
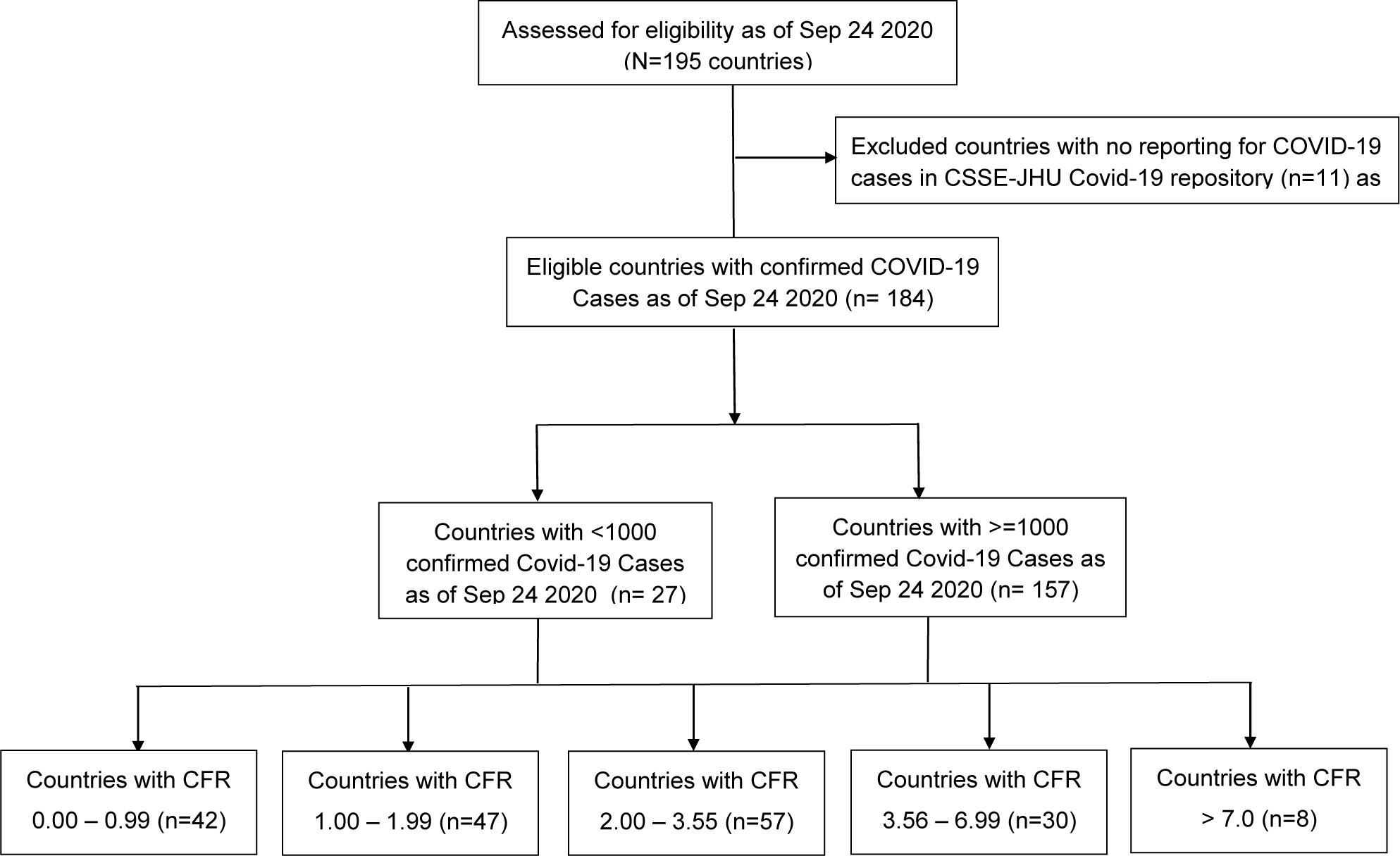
Study inclusion diagram as of 24 September 2020. 184 countries with at least one confirmed case of COVID-19 were eligible for this analysis. 157/184 countries had reported more than 1000 confirmed cases of COVID-19.

**Figure 2.A:**
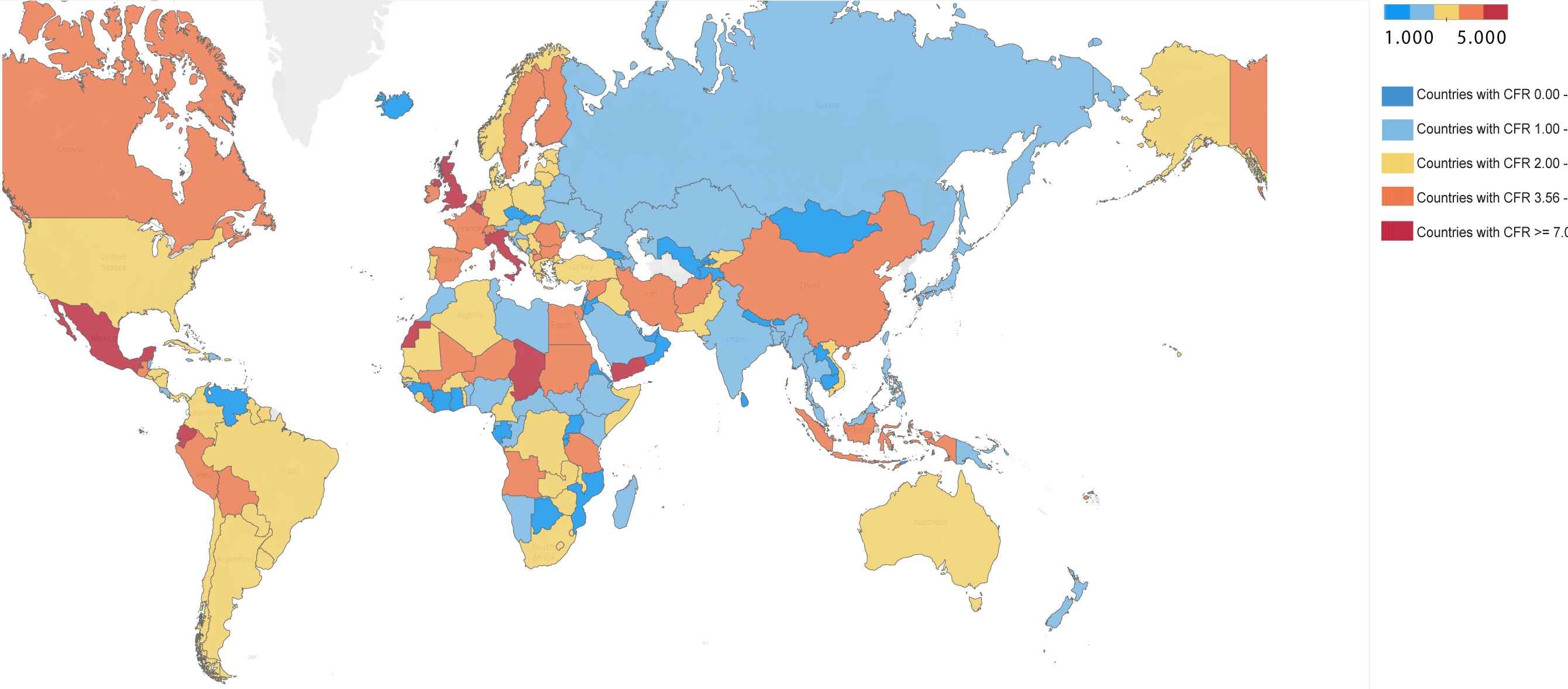
Global Distribution of COVID-19 case fatality ratio on September 24, 2020. World CFR on 24^th^ September was 3.06. 43 /157 countries report a CFR higher than the world CFR of 3.06

Unsupervised k-means clustering of 157 countries by their DP100K identified five clusters of countries (Supplementary Figure 1.A, Supplementary Table 5), A, B, C, D and E and hereafter referred to as country groups A, B, C, D and E. Optimal ***k*** for clustering was concurrent by CH-index in Tableau (Supplementary Table 6) as well as an elbow plot which showed a disruption of bent of the elbow curve between ***k*=5-6** (Supplementary Figure 2). Cluster separation was confirmed by Wilcoxon Signed Rank test which was significant for all five country groups at an alpha of 0.05 (Supplementary Figure 3, Supplementary Table 7).

Figure 2.B shows 157 world countries distributed by their country groups identified using ***k***-means clustering. Table 1 shows the number of countries, cases, deaths, CFR with 95%CI, DP100K, centroid DP100K, centroid CFR, and 10-day mean change for clusters A to E. Supplementary Table 4 shows the CFR with 95% CI and 10-day mean change in CFR for individual countries in each country group.

**Table 1:** The country group wise Case Fatality Ratio, Cases and Death on September 24, 2020 and Daily Mean Percentage Change (Sep 15 to Sep 24, 2020) in CFR. * Rounded to two decimal points

| Groups | No. of Countries | Cases | Deaths | CFR on Sep 24 2020 (95% CI*) | DP100K on Sep 24 2020 | Centroid DP100K | Centroid CFR | Starting date of COVID-19 Pandemic | Daily Mean % change from Sep 15 –Sep 24 2020 |
| --- | --- | --- | --- | --- | --- | --- | --- | --- | --- |
| **World** | 184 | 32140504 | 981792 | 3.06 (3.05-3.07) | 12.85 | NA | NA | 31-Dec | -0.11 |
| **Countries > 1000 cases** | 157 | 32133634 | 981664 | 3.06 (3.05-3.07) | 13.08 | NA | NA | 31-Dec | -0.11 |
| **Group A** | 6 | 102583 | 2573 | 2.51 (2.42-2.61) | 0.9 | 0.1 | 0.968 | 22-Jan | 0.09 |
| **Group B** | 26 | 362228 | 8253 | 2.28 (2.23-2.33) | 0.39 | 0.503 | 1.494 | 31-Dec | -0.05 |
| **Group C** | 36 | 1301476 | 22429 | 1.73 (1.71-1.75) | 2.05 | 1.816 | 1.961 | 22-Jan | -0.02 |
| **Group D** | 49 | 10209256 | 178686 | 1.76 (1.75-1.76) | 6.95 | 7.395 | 1.636 | 27-Jan | -0.04 |
| **Group E** | 40 | 20158091 | 769723 | 3.82 (3.82-3.83) | 52.87 | 36.303 | 3.589 | 22-Jan | -0.12 |

**Figure 2.B:**
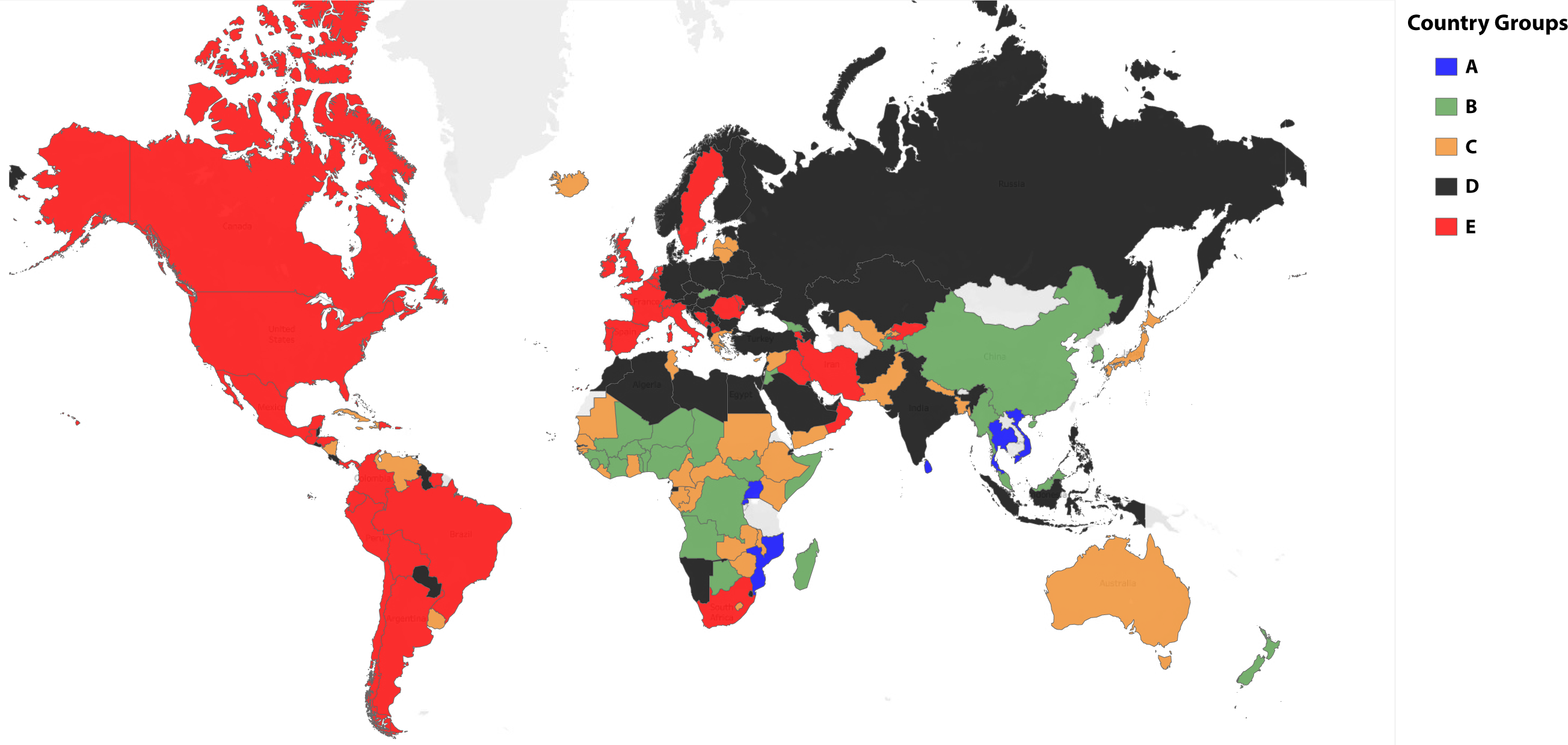
157 world countries with at least 1000 confirmed covid-19 countries distributed by their country groups identified using ***k***-means clustering

Figure 3 shows the day-wise time trend of CFR for the whole world, countries with case load >/= 1000 and country groups A to E and shows that the case fatality ratio of COVID-19 has been increasing over time in all these clusters since the beginning of the pandemic, followed by a plateau and then a downward trend. The CFR on September 24, 2020 for whole world, countries with >/= 1000 cases, Group A, Group B, Group C, Group D, and Group E were 3.06% (95% CI 3.057-3.07), 1.73 (95% CI 1.71-1.75), 2.51 (95% CI 2.42-2.61), 2.28 (95% CI 2.23-2.33), 1.73 (95% CI 1.71-1.75), 1.76 (95% CI 1.75-1.76), and 3.82 (95% CI 3.82-3.83), respectively (Table 1). Forty-three countries (1 in Group A, 6 in Group B, 6 in Group C, 9 in Group D and 21 in Group E) had a CFR higher than that of world CFR of 3.06. Consistent with this data, the CFR for Group E has tracked above the world average at all time points and this group, while accounting for only 19% of the world population, contributes to 63% of COVID-19 cases and 78.8% of COVID-19 deaths reported worldwide (Supplementary Figure 4 & Supplementary Table 8).

**Figure 3:**
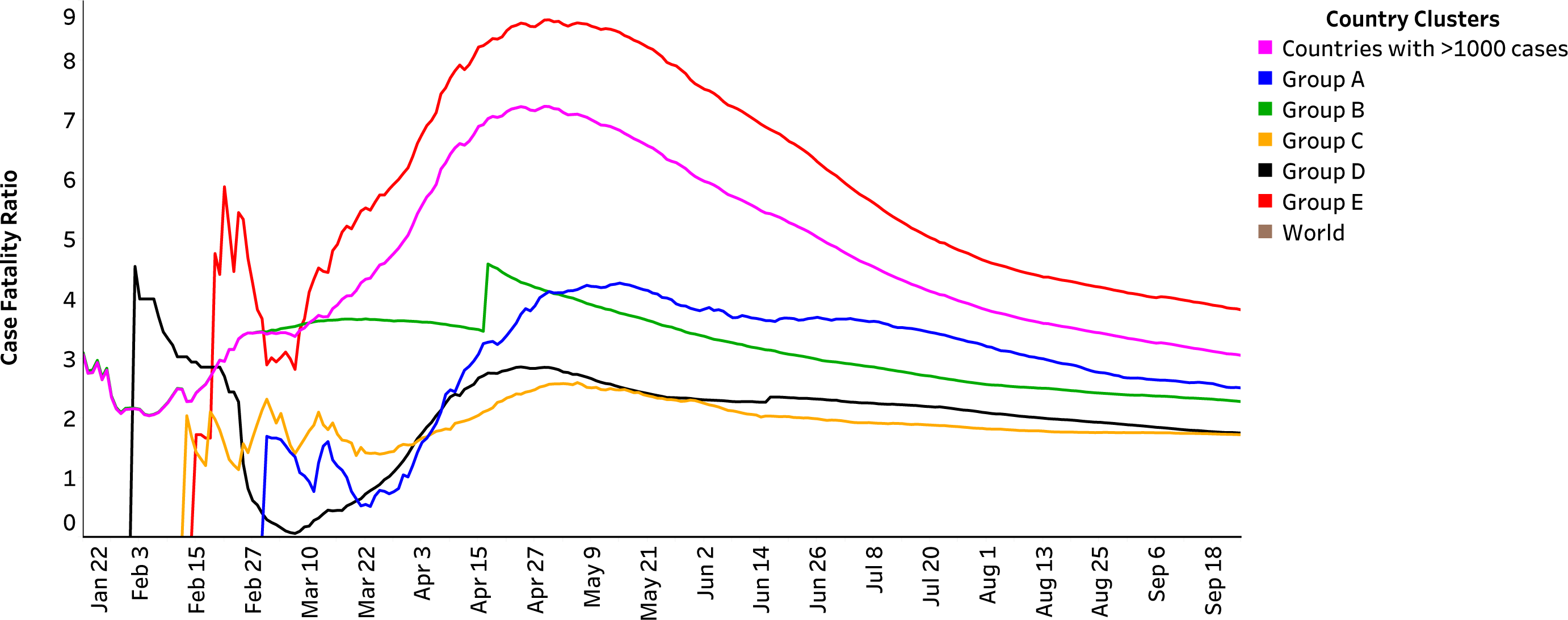
Case Fatality Ratio of Covid-19 pandemic across the world over time. Time trends of case fatality ratio for the whole world, included countries and clusters 1-5. X-axis indicates date and Y-axis indicates case fatality ratio. Each line depicts a Country Group. Purple line is the CFR over time for countries with atleast 1000 confirmed cases of COVID-19

Figures 4A, 4B, 4C, 4D and 4E depict the time trends of CFR in individual countries in five country groups along with the group CFR. For countries with highest DP100K (Group E, Figure 4E), the general trend is increasing CFR over time, followed by plateauing from late April 2020. As expected most countries show marked fluctuations in the day-wise CFR in the beginning of the pandemic because of the low number of cases and deaths. Curiously the CFR for China shows far less fluctuation in the beginning of the pandemic. Similarly, CFR for country groups A-D (Figures 4A-D respectively) also show an increasing trend over time with marked fluctuations within each country, especially in the beginning of the pandemic.

**Figure 4.**
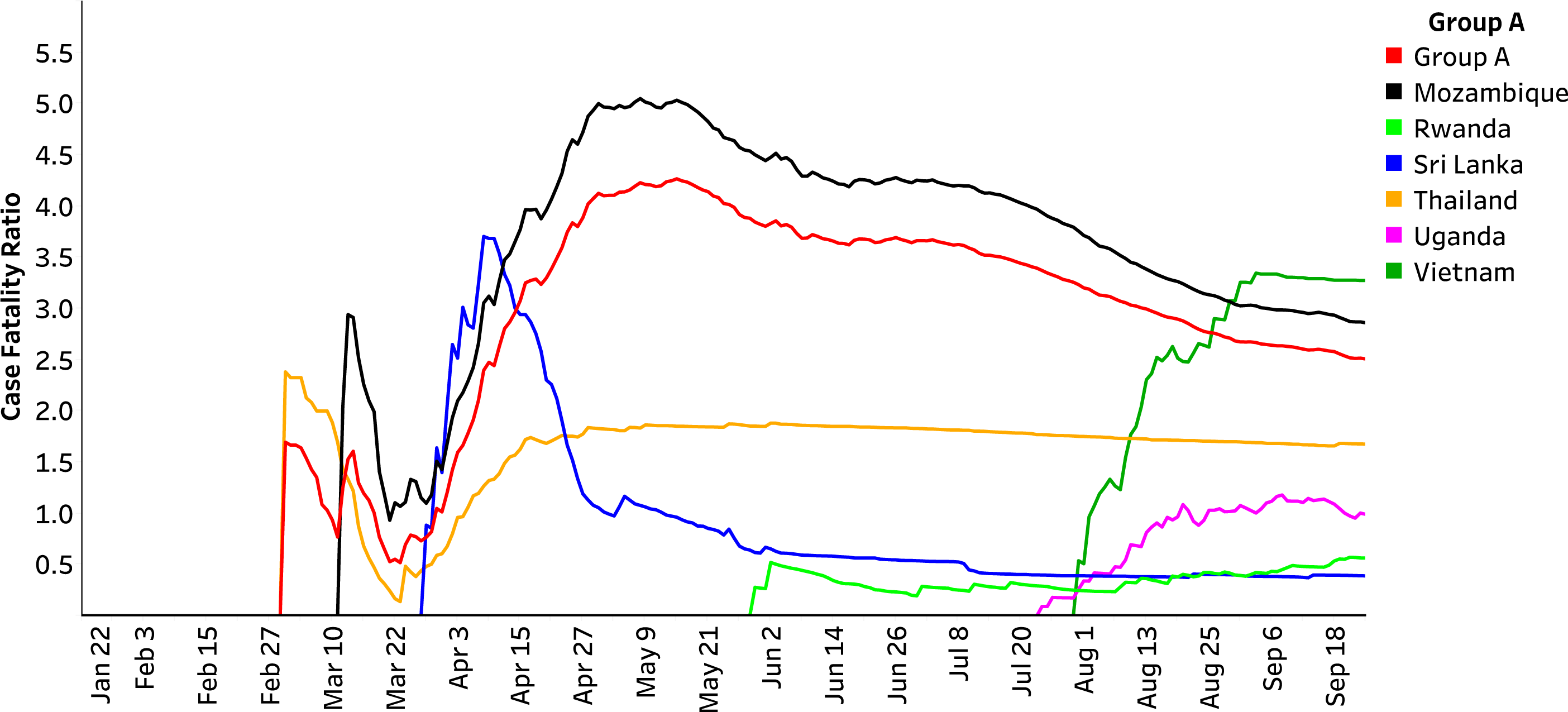
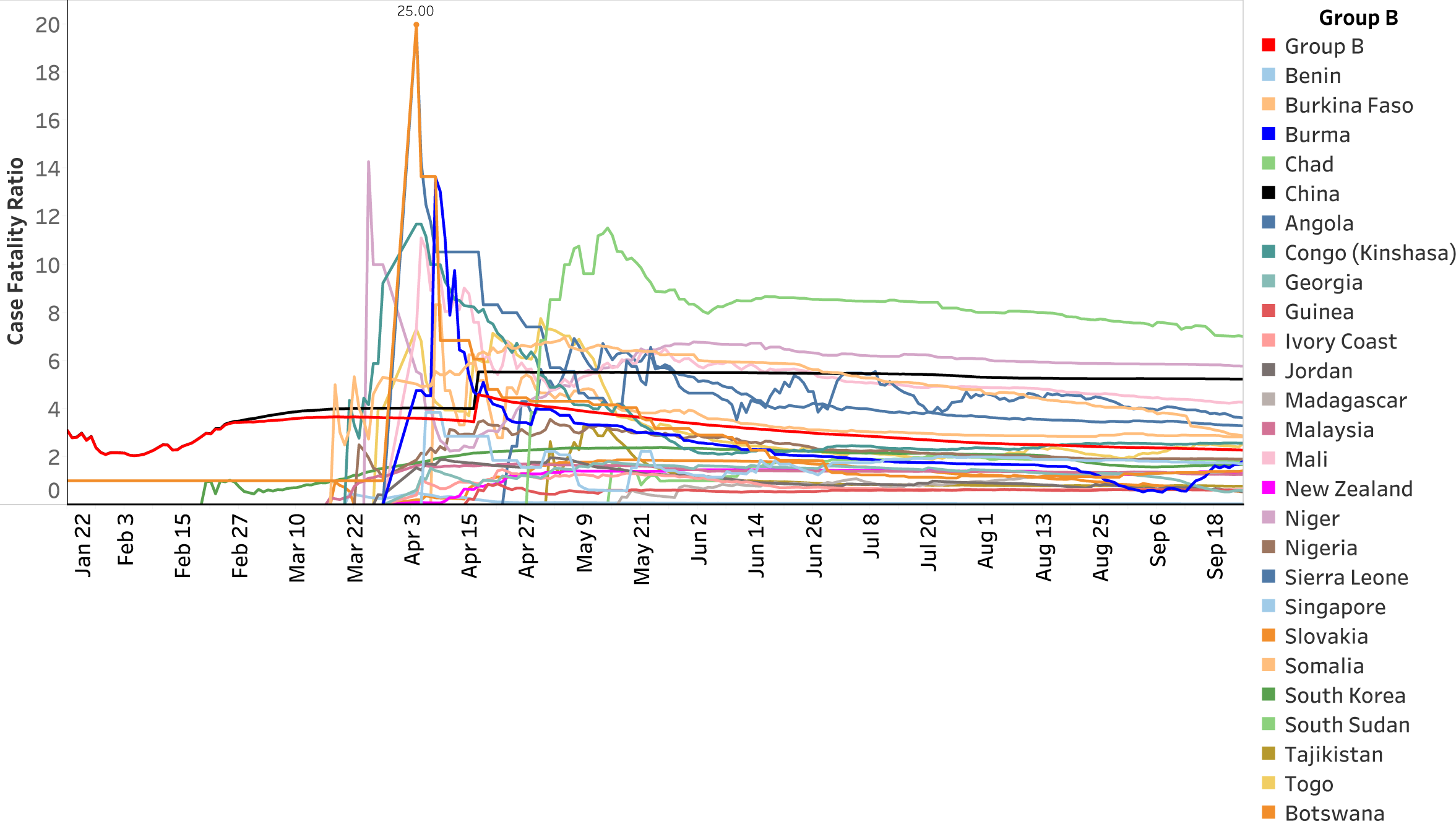
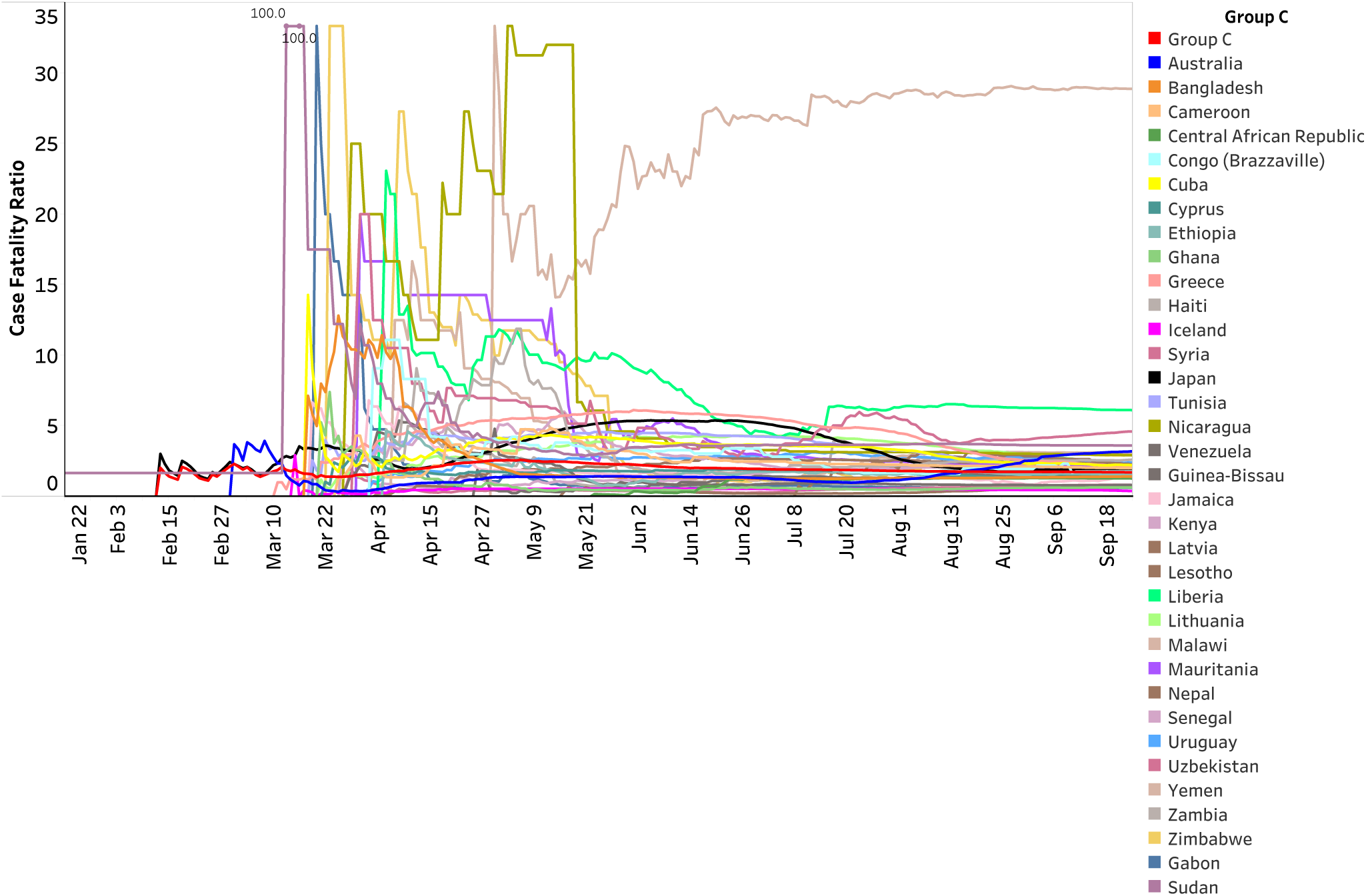
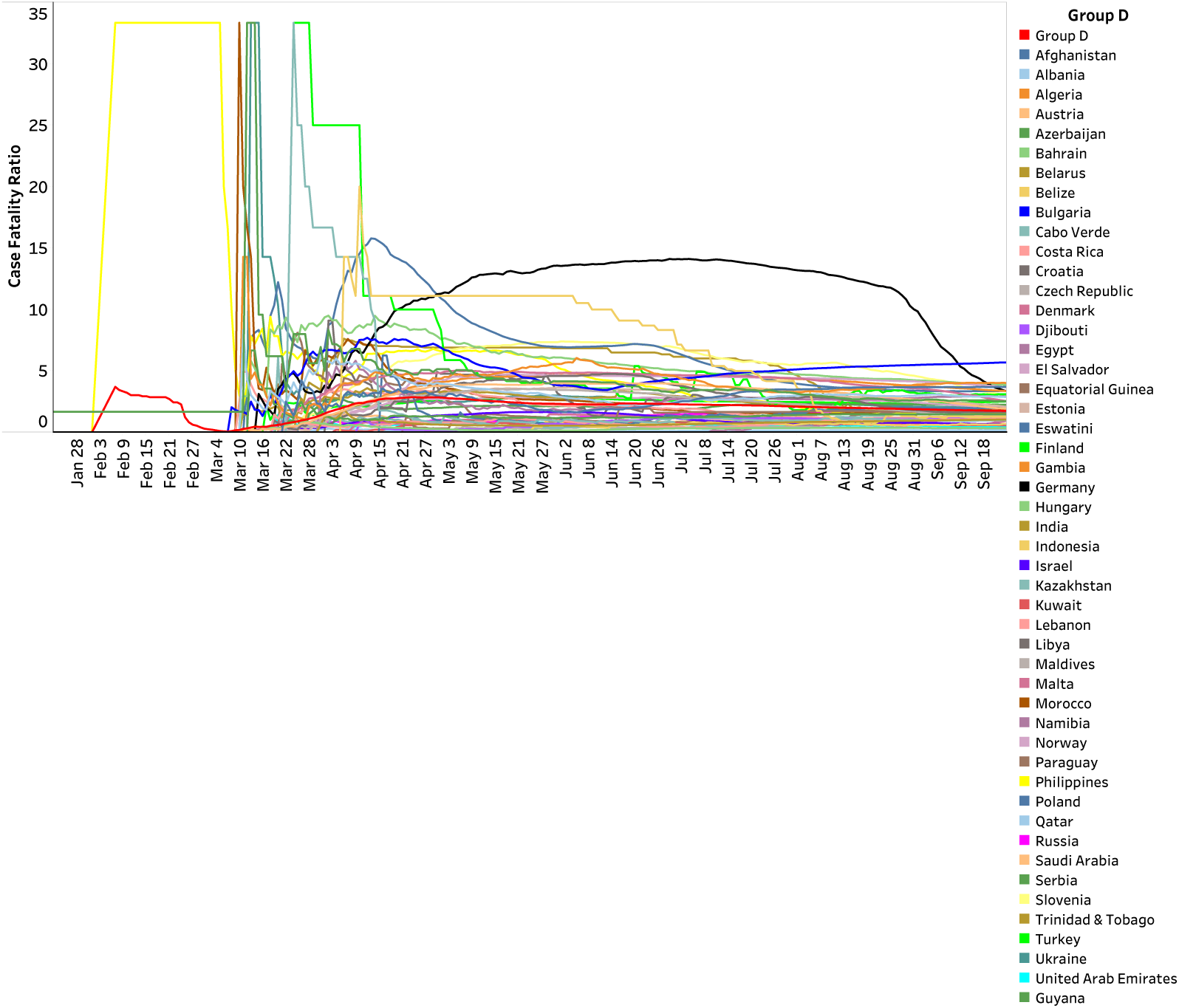
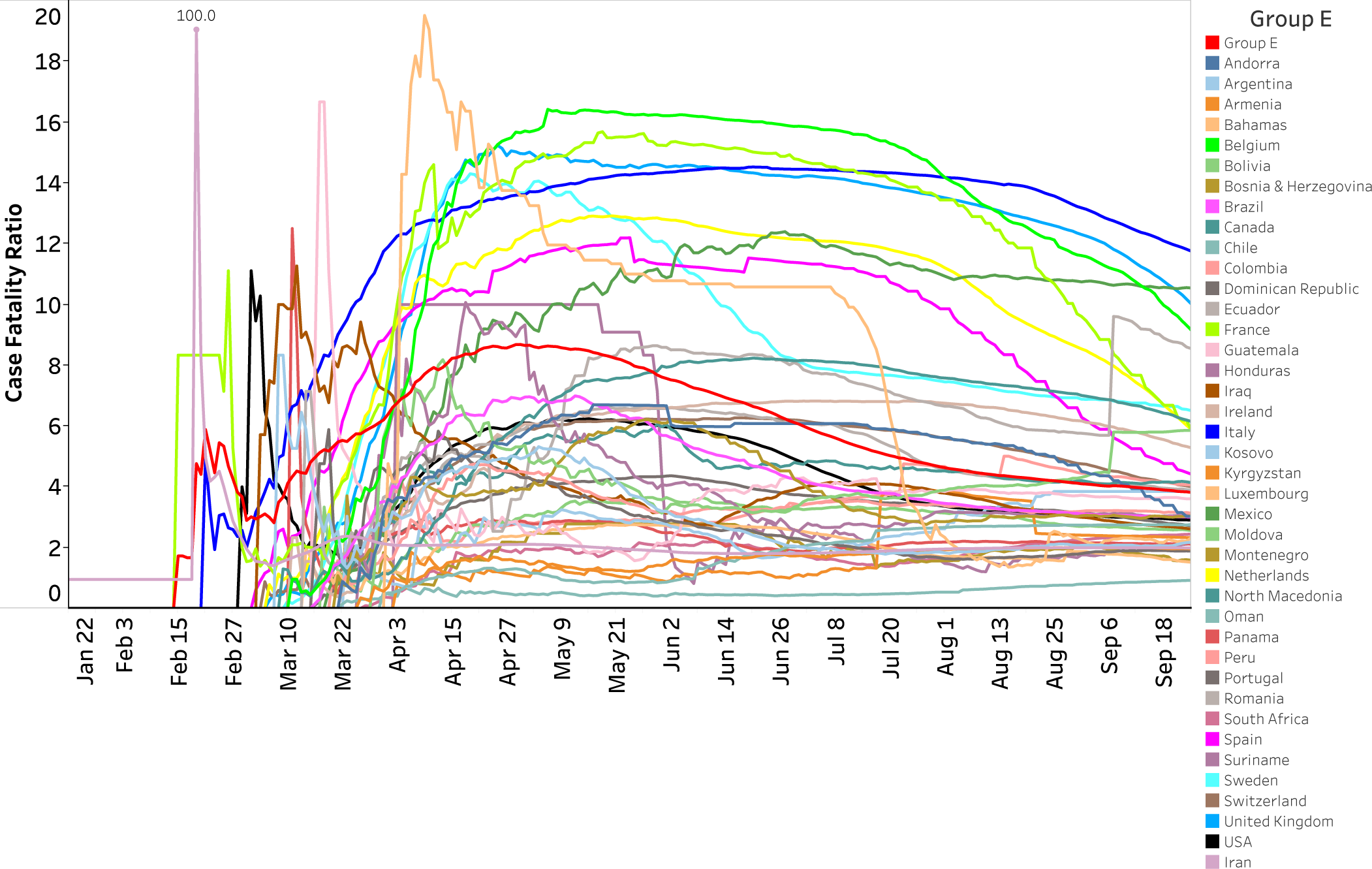
**A-E:** Time trends of case fatality ratio in Country Groups and individual countries in Group A (Figure 4A), Group B (Figure 4B), Group C (Figure 4C), Group D (Figure 4D) and Group E (Figure 4E). X-axis indicates date and Y-axis indicates case fatality ratio. Red line in each Figure is the CFR for the Group.

Supplementary Figure 5 shows the trend of CFR on the cutoff date as a function of case-load in 24 countries with more than 50,000 cases and DP100K >=20, and 3 additional countries with high case loads. The general pattern is that of increasing CFR with increasing case load. One country (China, Supplementary Figure 5 Inset A) shows low stable CFR followed by a sharp increase due to late reporting of previously unreported deaths in this country. The overall pattern suggests that CFR increases with case load in many countries as the healthcare system, especially intensive care, becomes overwhelmed (Supplementary Figure 5 Inset B. The United States has the maximum number of cases but has lower CFR than many countries with far fewer cases. This could be due to widespread testing and detection of many more patients with mild or asymptomatic disease but could also be due to the good quality healthcare infrastructure in United States.

### Correlation of CFR with Country-level Indices

Supplementary Table 9 shows the independent association of country-level health, human development, demographic, economic development and mobility indices with CFR in countries with case load >/= 1000 using weighted multiple linear regression. Multicollinearity assumption testing using Variance Inflation Factor (VIF) identified Infant Mortality rate/100,000 live births as being collinear with other indicators and was removed from the analysis. We also excluded smoking prevalence due to missing values from 30 countries. Our final model suggests that GDP per capita (β=-0.483, p=0.000), hospital beds per population (β=-0.372, p<0.001), mortality from air pollution (β=-0.487, p=0.003) and population density (β=-0.570, p< 0.000) were significantly negatively associated while maternal mortality ratio (β=0.431, p=0.000) and age (β=0.635, p<0.000) were positively associated with CFR. Of note, this model was able to account for much of the observed variation in CFR (R^2^=0.46).

## DISCUSSION

Our time series analysis using open source data suggests that the case fatality ratio of COVID-19 pandemic has increased over the period of its emergence in most countries and is currently at 3.06% for the aggregate of world data. The CFR of COVID-19 is twice that of the corresponding CFR (0.4-1.5%) reported by the World Health Organization for the H1N1 influenza pandemic [12]. The high CFR suggests that COVID-19 is a severe disease with important public health ramifications. The CFR has increased in countries with high number of cases as well as in those with lower case load. The gradually increasing CFR followed by plateau could be a manifestation of the long duration (2-8 weeks) [13] for COVID-19 related deaths to manifest in countries which have experienced rapid increase in case load in the past few months. Testing for SARS-CoV-2 has become more widely available and, in many countries, is now being offered to patients with less severe disease. This would be expected to increase the denominator and bring down the CFR and this may be the reason for the plateau that is becoming apparent in some countries and country groups.

As expected, the CFR in various countries and groups of countries is variable. This is likely because of the imbalanced distribution of factors like age and comorbidity status that impact COVID-19 related mortality and also because of variable availability of advanced intensive care infrastructure. It is also possible that unknown host or pathogen related factors are contributing to the variability in CFR. Because patient level data were not available to adjust potential confounding, we have not formally, statistically compared the observed CFR between various countries or clusters of countries. Our analysis suggests that there is a significant positive correlation between the country-level COVID-19 mortality burden as estimated by deaths per 100000 population (DP100K) and CFR (Supplementary Figure 1.B), but we are unable to decipher the presence or predominant direction of causality in this association. It is possible that CFR could contribute to DP100K, DP100K could contribute to CFR because of overwhelmed health infrastructure or unknown variable(s) could contribute to both in the same direction.

Other analyses have reported various figures for CFR of COVID-19 ranging from less than 1% to more than 10% [7, 14, 15, 16]. Some of these analyses have reported CFR after accounting for time-lag and other adjustments [17, 18, 19, 20]. It could be argued that the number of asymptomatic infected individuals is much larger than the current number of known infections and hence the CFR of COVID-19 is much lower than is estimated from so-called ‘naïve’ ratio that we have reported. While it might be true that the number of infected individuals is larger and the CFR among ‘universal set’ of infected individuals (the so-called infection mortality ratio) is lower, we would argue that the estimate of greatest interest from a clinical and health infrastructure perspective is the fraction of deaths among those individuals who are symptomatic. We would further argue that community level testing of asymptomatic individuals in the ‘screening’ mode and using that number to calculate CFR de-emphasizes the actual severity of the clinical disease.

One particular strength of our analysis is the relatively uniform definition of COVID-19 infection in this database, with a large majority being diagnosed by a single method of polymerase chain reaction. If and when serological tests enter widespread use and are used to diagnose COVID-19, the analysis of CFR will become more challenging. Our analysis of CFR as a time trend within countries is robust because the demographic and other variables within a country remain stable over the period of this analysis.

Our analysis in countries with heaviest COVID-19 involvement suggests a pattern of increasing CFR with increasing case load. This is likely due to high stress on the acute care infrastructure as cases increase. Countries with lesser case load at present should anticipate this pattern in the near future. This inference is further strengthened by the significant inverse association of hospital beds per unit population with CFR in the country-level analysis.

Our country-level analysis suggests that higher resources (higher GDP per capita and hospitals beds per population) are associated with lower CFR, which attests to the imperative of supporting countries with lesser resources. Higher age is also independently associated with CFR which could explain the relatively high fatalities in some European countries. An enigmatic inverse association in our analysis is with population density. Many countries with high density are also the ones with lesser resources and the lower CFR and deaths in these countries, relative to their population sizes, is as yet unexplained.

There are some limitations of our analysis. We did not account for time- or severity-dependent reporting of cases and also for the time-lag in outcomes. It is possible that COVID-19-related deaths have been underreported in some parts of the world and that the actual CFR is higher than our estimates. We did not have access to patient-level data and data of such granularity may never become available for such a large number of cases and deaths. Therefore, we have been unable to provide ‘adjusted’ CFR after accounting for the distribution of variables. The correlation between CFR and health and social parameters could possibly be masked or confounded because the latter were available at the country-level with likely intra-country variation.

In summary, our analysis confirms that COVID-19 is a severe disease in a considerable minority of patients, its CFR has gradually increased in many parts of the world, and, given the large number of cases, is likely to result in high absolute number of deaths unless an effective vaccine or treatment is developed. Our analysis also indicates that the herd immunity argument is likely inappropriate and will possibly result in a high number of fatalities. Governments and public health authorities should consider the CFR among clinically presenting cases and plan acute care capacity accordingly.

## Data Availability

All the raw data used in this analysis
is available as supplementary files

## Supplementary Figures

**Supplementary Figure 1: A) Unsupervised clustering of countries by DP100K of the population**. Log transformed Case Fatality Ratio (CFR) (y-axis) plotted against log transformed Death per 100,000 of the population (DP100K). Each data point represents the CFR and DP100K for a country. Clusters are colour coded. **B) Linear regression of DP100K vs CFR for 157 Countries in five cluster group countries**. Our model generated an R-squared of 0.12748 with a standard error of 0.12748 and p-value < 0.0001

**Supplementary Figure 2:** Elbow plot to identify optimal *k* for k-means clustering of 157 countries by their DP100K.

**Supplementary Figure 3:** Violin plots for CFR in five country groups. We excluded CFR for Yemen as it was an outlier and skewing the results.

**Supplementary Figure 4:** Percentage distribution of total cases, deaths and active cases across countries and clusters with respect to their population.

**Supplementary Figure 5: CFR as a function of case load**. Cumulative cases for a country are plotted on x axis against the CFR for those cases on y axis. Wherever data is missing between successive two data points, a rolling average of those two data points is used to join them with a straight line.

**Supplementary Figure 5 (Inset A):** CFR as a function of Case Load for countries upto 150,000 cases.

**Supplementary Figure 5 (Inset_B):** CFR as a function of Case Load for countries upto 500,000 cases.

## Supplementary Tables

**Supplementary Table 1:** Raw data downloaded from the CSSE-JHU github repository on 25^th^ September 2020 and used for analysis

**Supplementary Table 2:** 10 country level indices for health, human development, population demographics and economic development downloaded from the world bank open data hub repository and the UNO data bank and used for analysis

**Supplementary Table 3:** Data points removed from visualization in figure 4 to enable better visualization of CFR trends over time.

**Supplementary Table 4:** CFR time trends, 95% CI levels and 10 day mean change for countries in the analysis

**Supplementary Table 5:** 157 countries clustered on the basis of their log DP100K

**Supplementary Table 6:** C-H index statistical Model for k-means clustering of 157 countries by their DP100K

**Supplementary Table 7:** Wilcoxon-rank test to identify differences in median CFR for all five country groups

**Supplementary Table 8:** Cases, deaths and active cases in five country groups and countries with less than 1000 cases as a percentage of the whole world.

