## Supplementary figures 1-5 for "WORLDWIDE CASE FATALITY RATIO OF COVID-19 OVER TIME"

Sfig1.A: Unsupervised clustering of countries by DP100K of the population

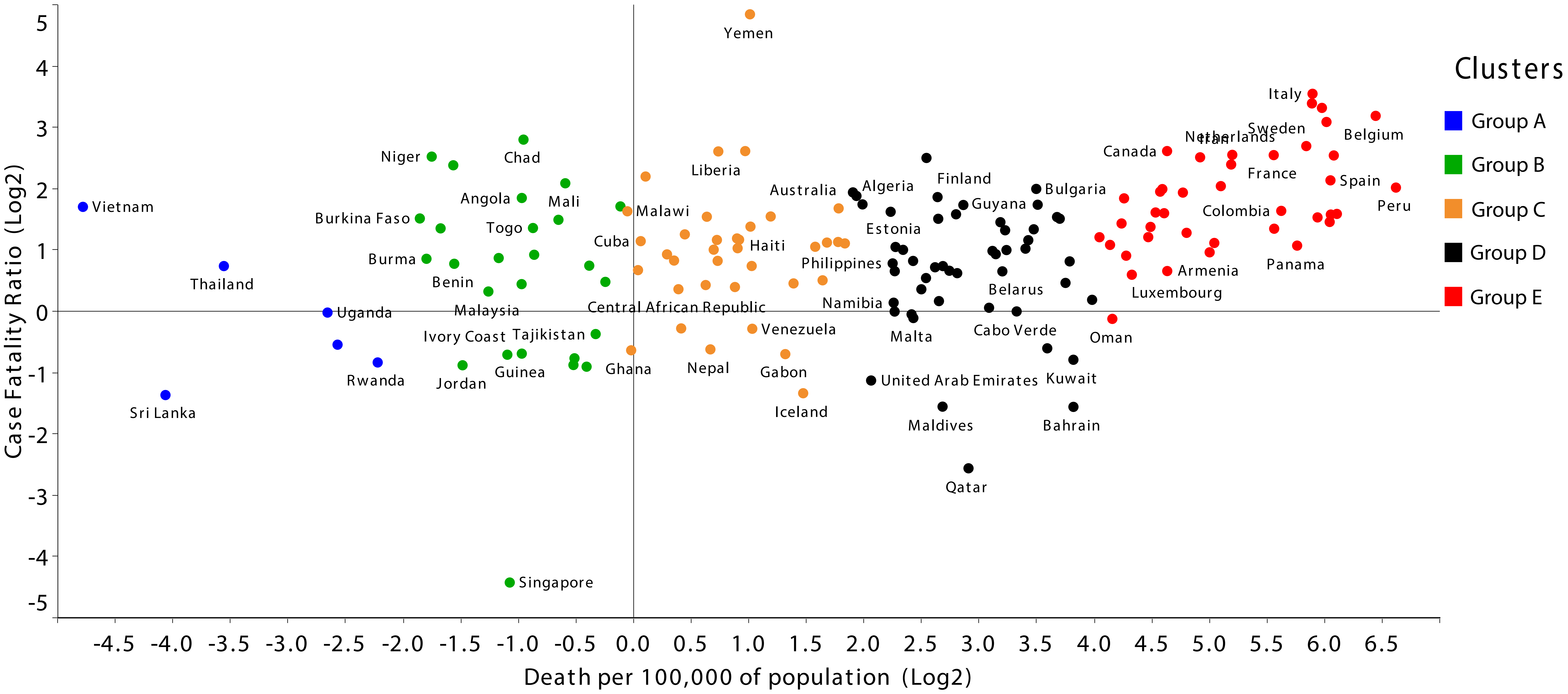


Log transformed Case Fatality Ratio (CFR) (y-axis) plotted against log transformed Death per 100,000 of the population (DP100K). Each data point represents the CFR and DP100K for a country. Clusters are colour coded

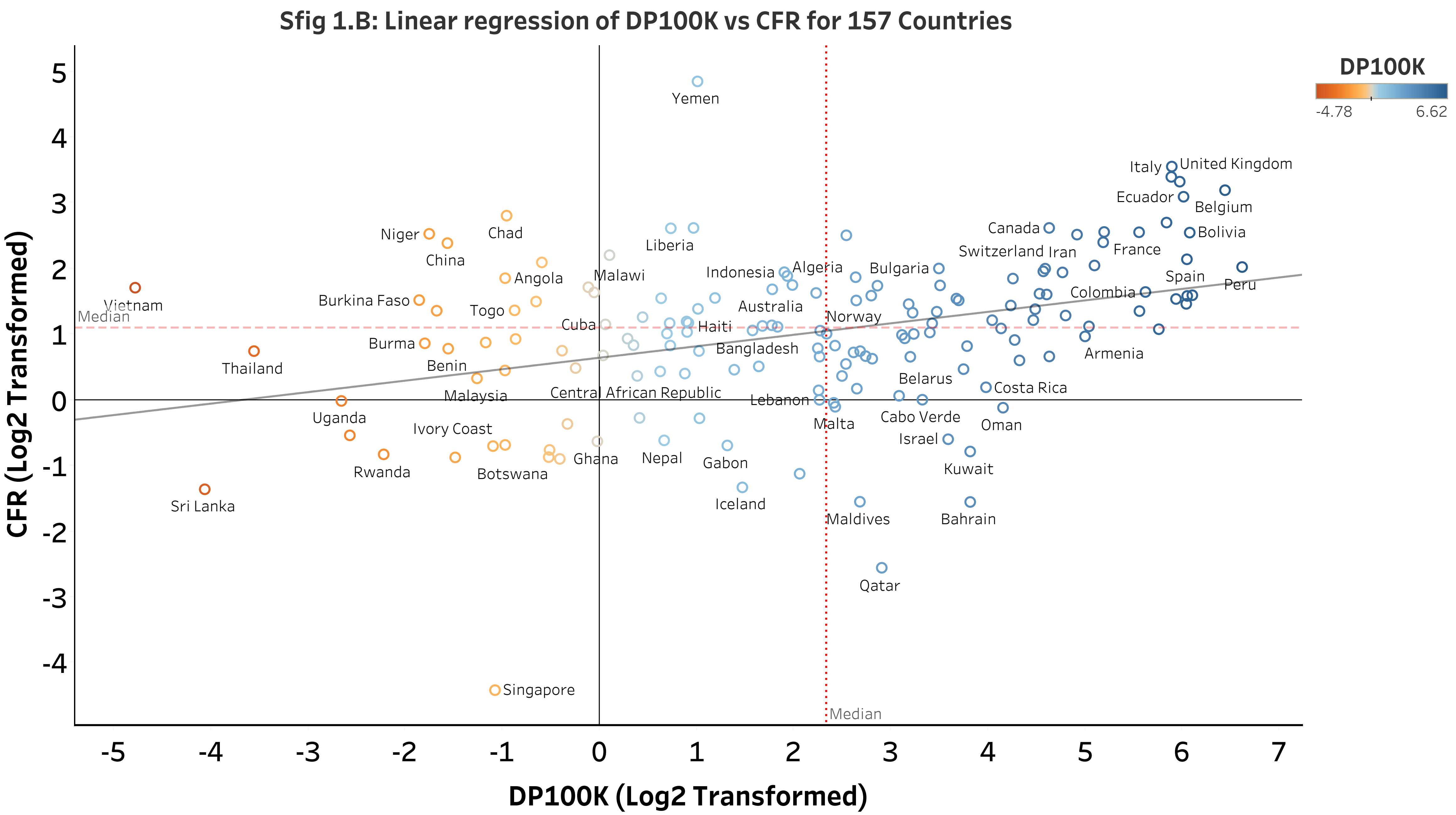
Sfig 1.B: Linear regression of DP100K vs CFR for 157 Countries
DP100K
-4.78
6.62
CFR (Log2 Transformed)
DP100K (Log2 Transformed)
Median
Median
Yemen
Italy
United Kingdom
Ecuador
Belgium
Canada
Bolivia
Switzerland
Iran
France
Chad
Niger
China
Liberia
Malawi
Indonesia
Algeria
Bulgaria
Spain
Peru
Angola
Colombia
Burkina Faso
Vietnam
Togo
Australia
Norway
Cuba
Haiti
Bangladesh
Burma
Armenia
Thailand
Benin
Belarus
Malaysia
Central African Republic
Costa Rica
Lebanon
Uganda
Cabo Verde
Malta
Oman
Ivory Coast
Israel
Rwanda
Ghana
Nepal
Gabon
Kuwait
Botswana
Sri Lanka
Iceland
Maldives
Bahrain
Qatar
Singapore

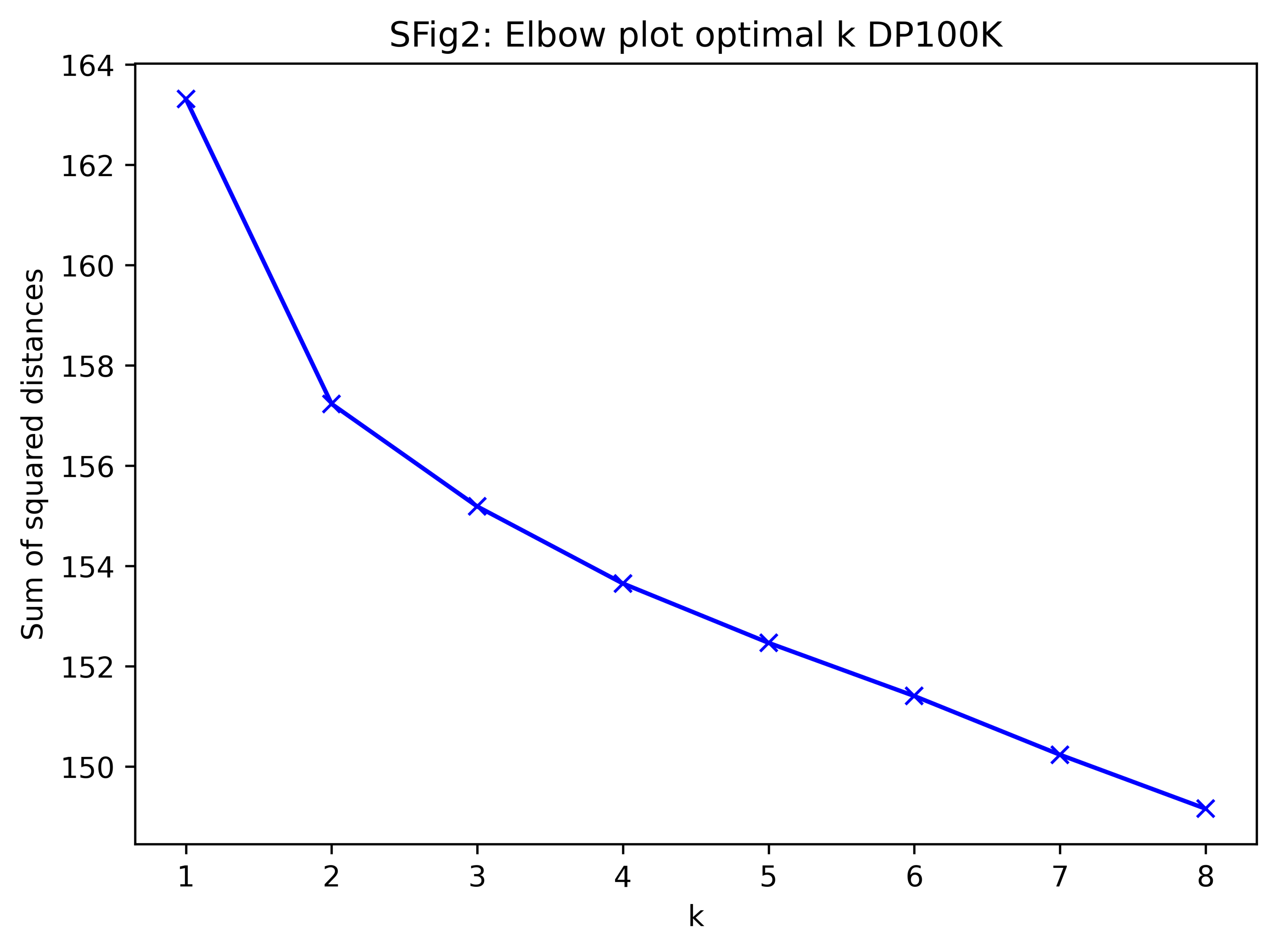
SFig2: Elbow plot optimal k DP100K
Sum of squared distances
164
162
160
158
156
154
152
150
1
2
3
4
5
6
7
8
k

#### Sfig3: Violin plots for CFR in five country groups

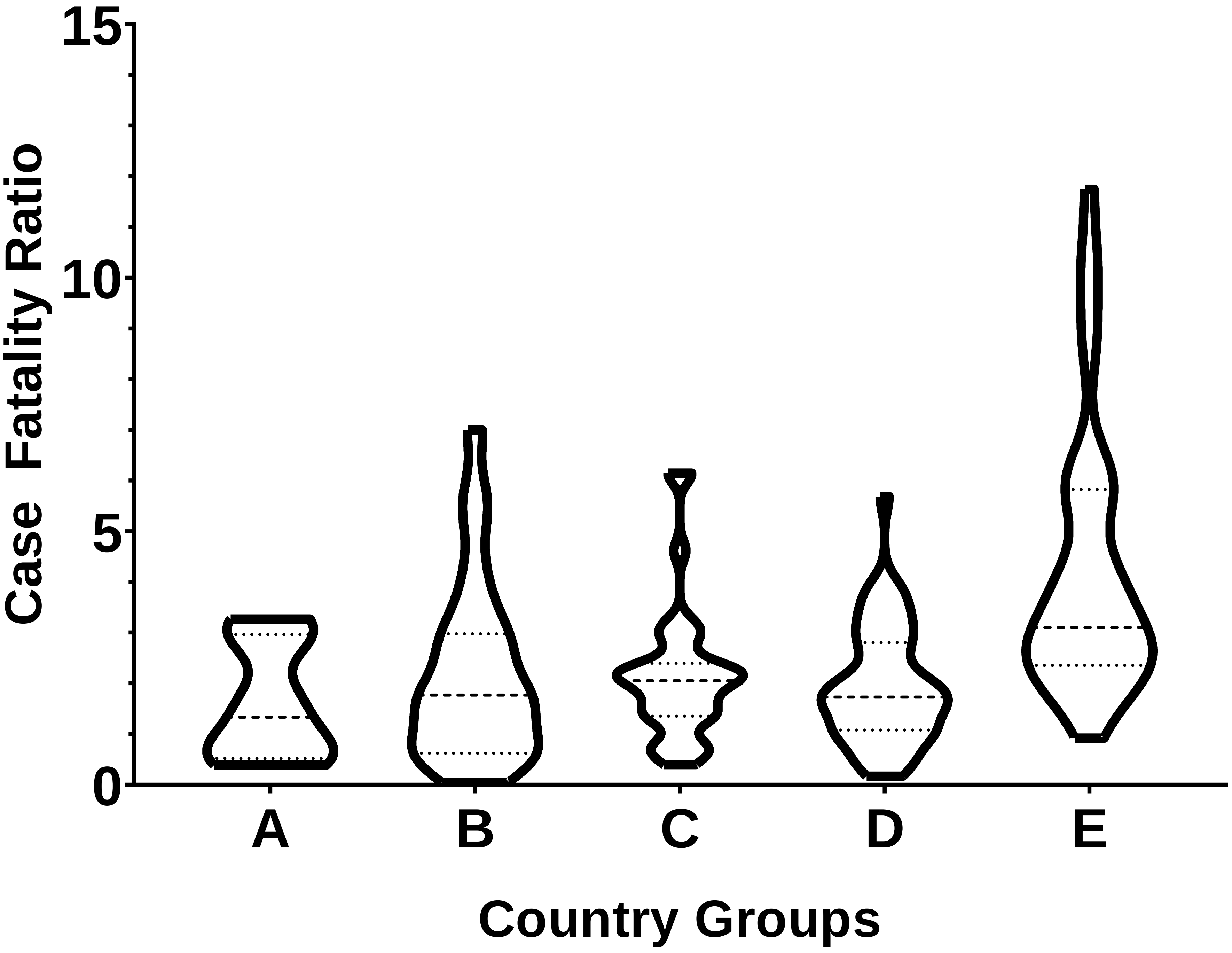

SFig 4: Percentage distribution of total cases, deaths and active cases across countries and clusters with respect to their population

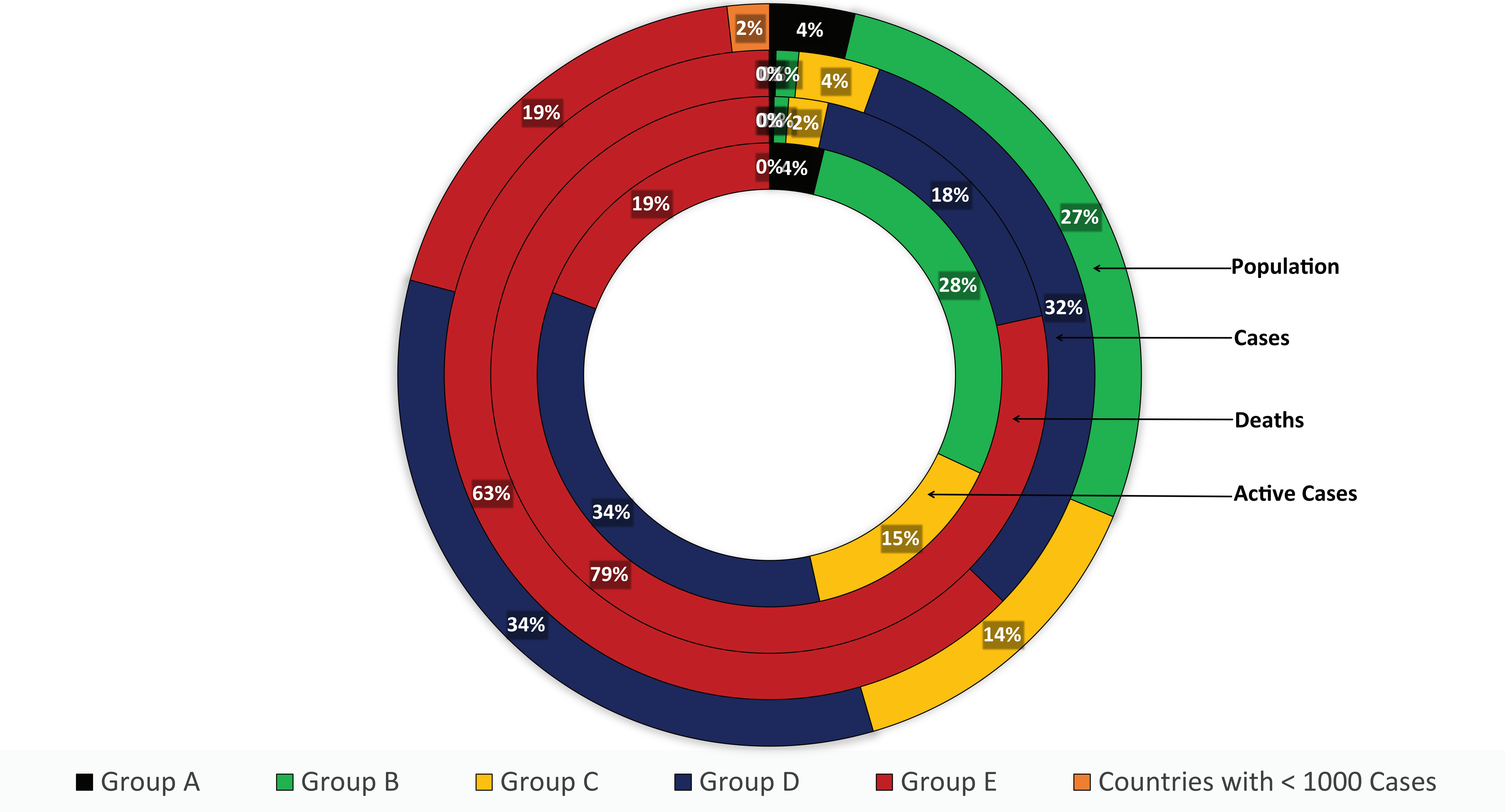

Sfig 5: CFR as a function of Case Load

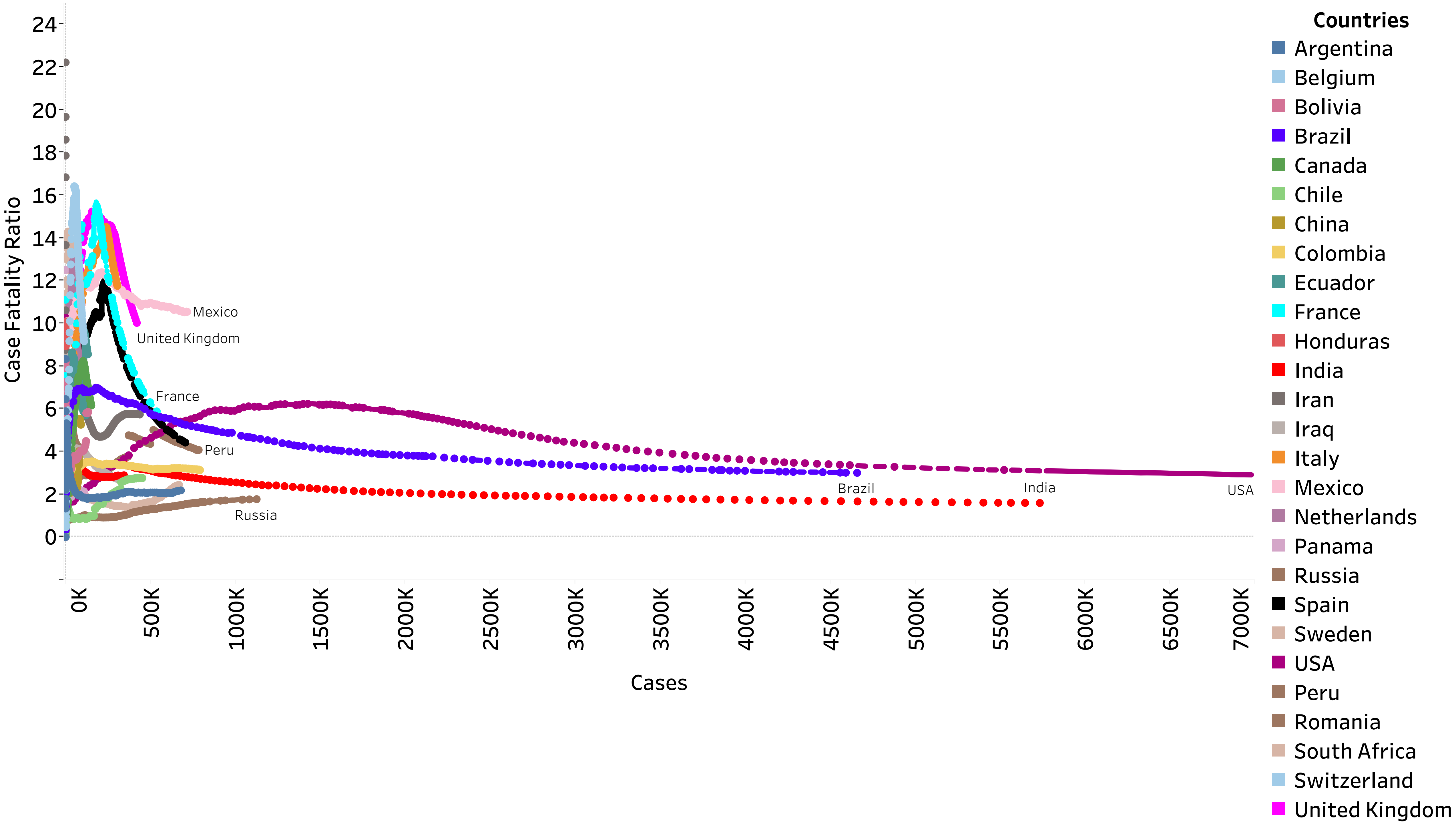

### Sfig 5 (Inset A): CFR as a function of Case Load for countries upto 150,000 cases

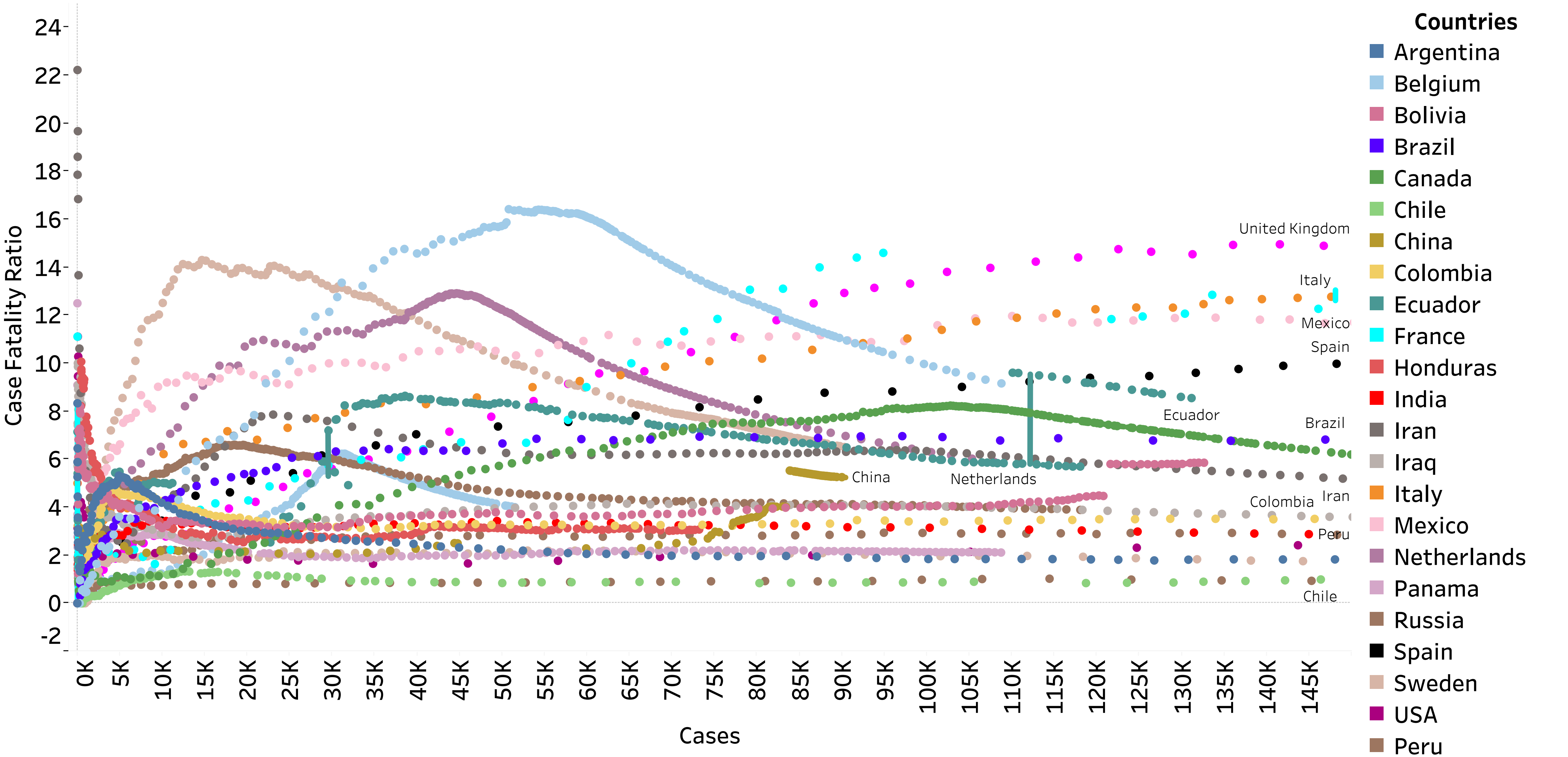

Sfig 5 (Inset B): CFR as a function of Case Load for countries upto 500,000 cases

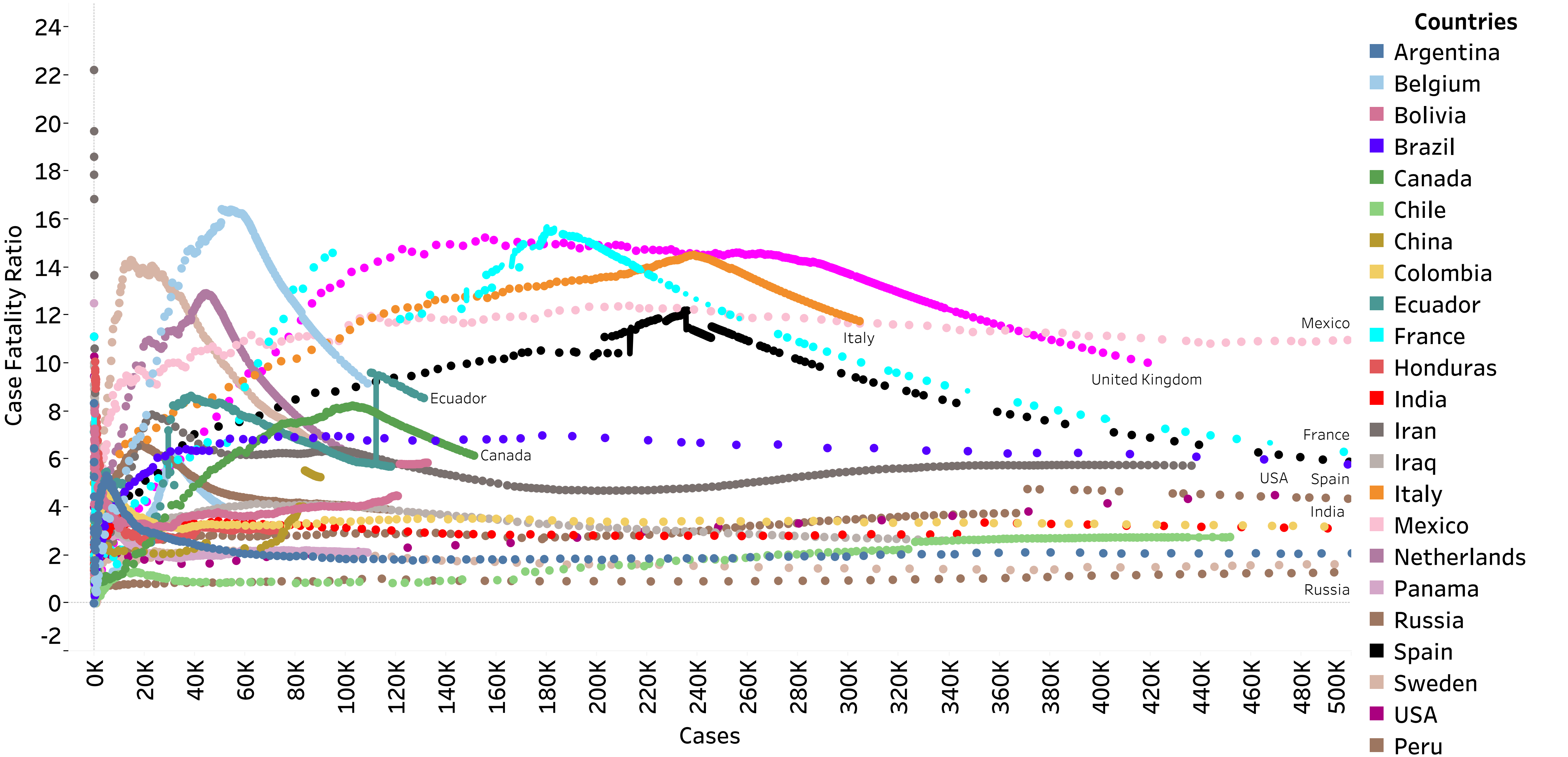
