## Supplementary Table 9 for "WORLDWIDE CASE FATALITY RATIO OF COVID-19 OVER TIME"

**Supplementary Table 9: Weighted multiple linear regression analysis with 10 country level indicators of health, human developement, economic developement, and population demographics. Two indicators (Infant mortality rate and smoking % per population were eliminated in colinearity analysis. Resulting model was evaluated with 8 indcators.**

**Model Summary**

| Model | R | R Square | Adjusted R Square | Std. Error of the Estimate | Change Statistics | | | | |
|---|---|---|---|---|---|---|---|---|---|
| | | | | | R Square Change | F Change | df1 | df2 | Sig. F Change |
| 1 | .678[a] | .460 | .428 | 37.35877 | .460 | 14.470 | 8 | 136 | .000 |

a. Predictors: (Constant), Median Age, Population Density / sqkm, % Mortality from CVD, cancer, diabetes or CRD (Age 30 - 70 ), Hospital Beds / 1K, MMR / 100K live births, GDP / Capita, Physicians / 1k, ASR Mortality air pollution / 100K

**ANOVA[a,b]**

| Model | | Sum of Squares | df | Mean Square | F | Sig. |
|---|---|---|---|---|---|---|
| 1 | Regression | 161558.766 | 8 | 20194.846 | 14.470 | .000[c] |
| | Residual | 189812.174 | 136 | 1395.678 | | |
| | Total | 351370.939 | 144 | | | |

a. Dependent Variable: CFR

b. Weighted Least Squares Regression - Weighted by Inv_Sesq

c. Predictors: (Constant), Median Age, Population Density / sqkm, % Mortality from CVD, cancer, diabetes or CRD (Age 30 - 70 ), Hospital Beds / 1K, MMR / 100K live births, GDP / Capita, Physicians / 1k, ASR Mortality air pollution / 100K

**Coefficients[a,b]**

| Model | | Unstandardized Coefficients | | Standardized Coefficients | t | Sig. | 95.0% Confidence Interval for B | | Collinearity Statistics | |
|---|---|---|---|---|---|---|---|---|---|---|
| | | B | Std. Error | Beta | | | Lower Bound | Upper Bound | Tolerance | VIF |
| 1 | (Constant) | -1.219 | 1.277 | | -.955 | .341 | -3.744 | 1.305 | | |
| | GDP / Capita | -2.712E-005 | .000 | -.483 | -4.617 | .000 | .000 | .000 | .363 | 2.756 |
| | MMR / 100K live births | .007 | .002 | .431 | 3.993 | .000 | .004 | .011 | .341 | 2.930 |
| | Hospital Beds / 1K | -.280 | .083 | -.372 | -3.370 | .001 | -.445 | -.116 | .326 | 3.070 |
| | Physicians / 1k | .224 | .152 | .168 | 1.471 | .144 | -.077 | .524 | .304 | 3.292 |
| | % Mortality from CVD, cancer, diabetes or CRD (Age 30 - 70 ) | -.028 | .039 | -.096 | -.714 | .476 | -.106 | .050 | .220 | 4.536 |
| | ASR Mortality air pollution / 100K | -.010 | .003 | -.487 | -3.065 | .003 | -.017 | -.004 | .157 | 6.357 |
| | Population Density / sqkm | .000 | .000 | -.570 | -6.572 | .000 | -.001 | .000 | .528 | 1.896 |
| | Median Age | .160 | .037 | .635 | 4.279 | .000 | .086 | .234 | .181 | 5.537 |

a. Dependent Variable: CFR

b. Weighted Least Squares Regression - Weighted by Inv_Sesq
